# A comprehensive phenome wide analysis of the role of neutrophils in health and disease

**DOI:** 10.1101/2025.02.06.25321790

**Authors:** K Fleming, N Cornish, EE Vincent, AD Mumford, B Amulic, K Burley

## Abstract

Neutrophil release of cytoplasmic granules containing antimicrobial agents is a critical component of innate immunity. Neutrophils are widely implicated in other disease responses yet the overall extent of the neutrophil contribution to human health and disease is incompletely characterized. To explore this further, we leveraged publicly available genetic data to conduct a Mendelian randomization phenome-wide association study (MR-PheWAS) of neutrophil traits and 14,983 outcomes.

Genetic proxies for neutrophil count, granularity, and serum myeloperoxidase were linked to 146 outcomes. Higher neutrophil count was associated with lower body weight, reduced obesity risk, and increased vascular activation markers but not with atherosclerosis. Elevated neutrophil count was robustly linked to Alzheimer’s disease and neutrophil granularity with gut microbiota abundance and dental pathology.

Our findings reveal the diverse roles of neutrophils extending beyond pathogen defense and underscore the potential for MR-PheWAS in identifying novel neutrophil-related pathophysiology.

**Summary sentence:** Neutrophils, essential for immunity, also influence other aspects of health, as shown in our MR-PheWAS study linking genetic proxies for neutrophil traits to 146 outcomes, including obesity, Alzheimer’s disease, endothelial activation markers and gut microbiota.

**VISUAL ABSTRACT:** 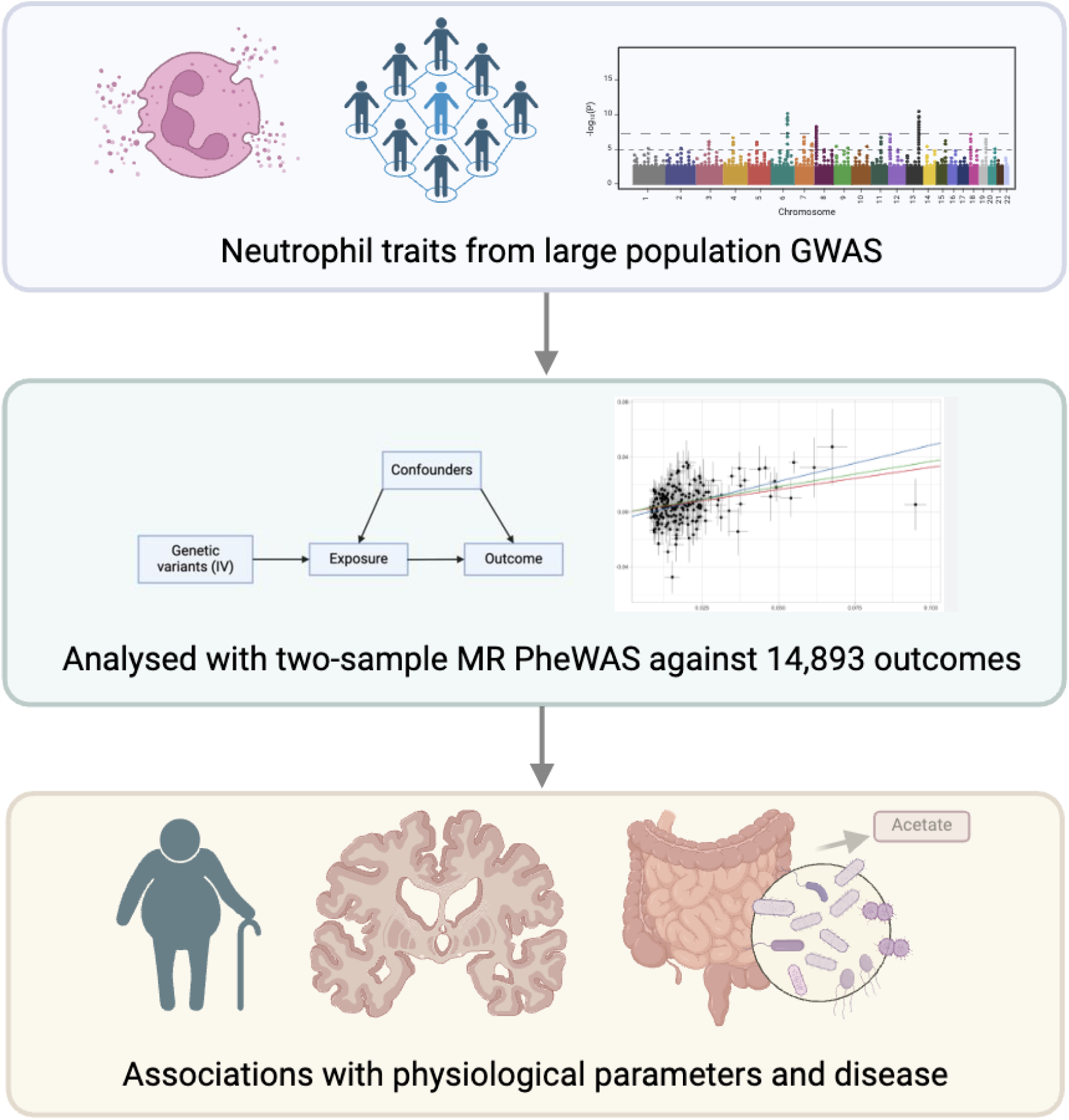

## INTRODUCTION

Neutrophils are the most abundant circulating leucocytes in human blood and function as essential mediators of the innate immune response against invading microorganisms. This function requires recruitment via chemokines to extravascular sites of infection where neutrophils rapidly contain bacterial and fungal proliferation by phagocytosis and the secretion of cytoplasmic granules packed with potent antimicrobial agents, including myeloperoxidase (MPO), lysozyme and defensins.(1) In keeping with this critical function, disorders associated with reduced neutrophil number and/or function are associated with immunodeficiency.

There is increasing recognition that neutrophils are also major cellular mediators of inflammation. This activity may contribute to tissue injury responses to infection and to the pathogenesis of autoimmune diseases such as lupus, inflammatory bowel disease and rheumatoid arthritis.(2–4) In addition, neutrophils have been implicated in both the development of atherosclerosis and amplification of cardiac injury post-myocardial infarction, with neutrophil count and serum MPO concentration correlating with adverse clinical outcomes.(5–7)

Many of the reported associations between traits such as neutrophil count and disease outcomes are observational only, thereby creating uncertainty whether neutrophils are causally implicated in pathology or alterations in neutrophil traits are the consequence of disease. Mendelian randomization (MR) is a well-established instrumental variable analysis that addresses some of the shortcomings of conventional observational studies by using genetic proxies for exposures, such as neutrophil traits, to evaluate causal relationships with disease outcomes.(8) In two-sample MR, summary-level data from genome wide association studies (GWAS) can be used, providing specific assumptions are met, to estimate causal effects without confounding from acquired risk factors (Figure 1A). MR can then be repeated across numerous outcome traits to systematically scan the phenome for associations with exposures in a hypothesis-free approach.

**Figure 1:**
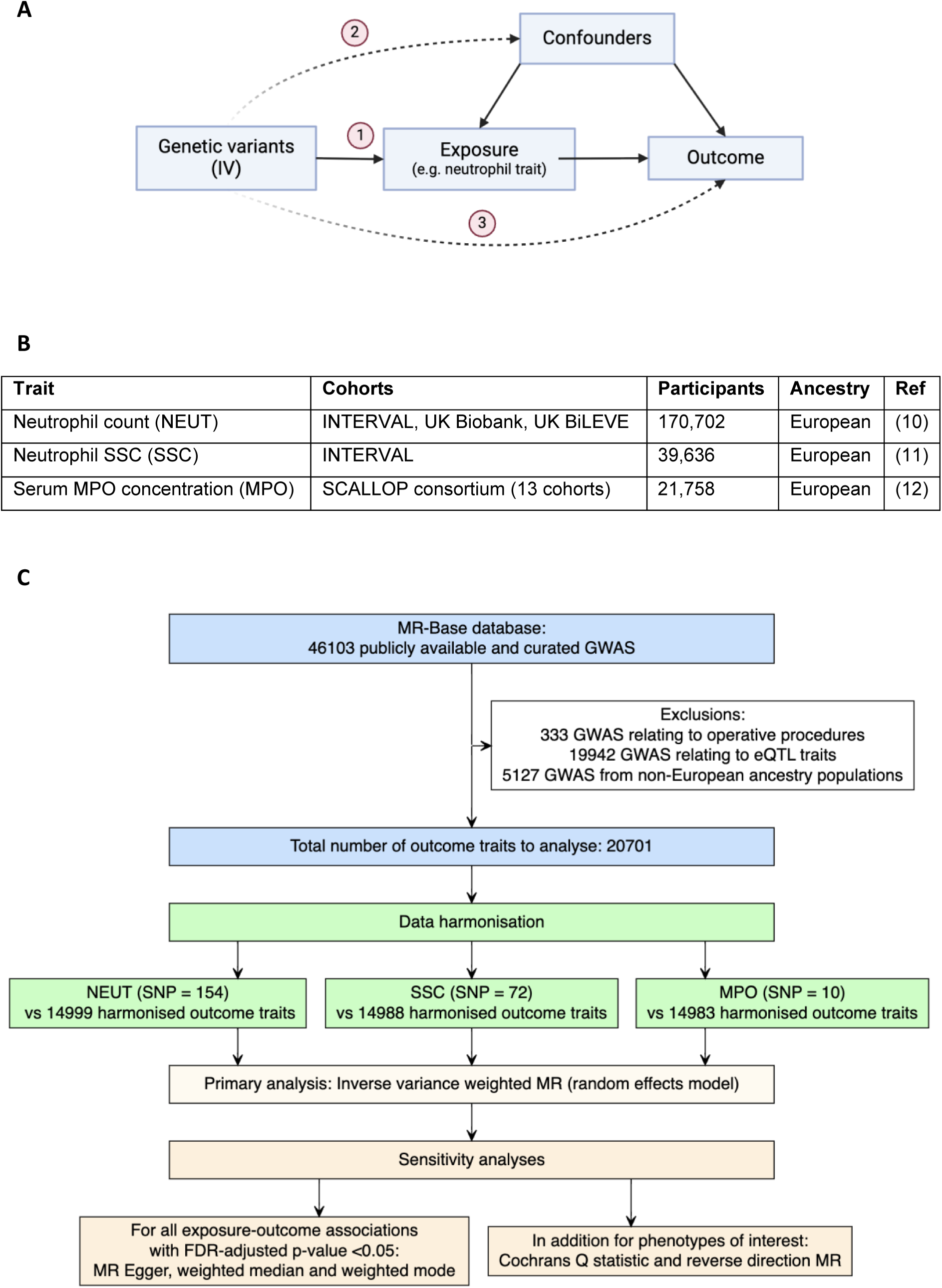
MR-PheWAS methodology. (A) Directed acyclic graph (DAG) depicting the principle of MR and underlying instrumental variable (IV) assumptions. Solid arrows indicate causal links, and dashed arrows indicate potential violations of MR assumptions (1–3). Assumption 1: The IV is associated with the exposure of interest (*relevance*), which can be assessed using the F-statistic. Assumption 2: The IV is associated with the outcome only through the exposure (*independence*). With control for population structure in the source GWAS, Mendel’s law implies that IVs will not be directly associated with confounders. Assumption 3: The IV influences the outcome only through the exposure and not through alternative pathways (*exclusion restriction*). Primary MR analyses typically assume no pleiotropy, however sensitivity analyses such as MR Egger calculate the effect estimate adjusted for any directional pleiotropy. (B) Details of GWAS from which exposure instruments were constructed. (C) MR-PheWAS workflow. Three separate PheWAS were undertaken to examine the causal effect of neutrophil traits (exposures) on a range of outcomes. IVs for the exposures were selected, data for these variants were extracted from outcome GWAS summary statistics and then harmonized to ensure that variant effects on the exposure and outcome corresponded to the same allele. Two-sample MR was undertaken, and results interrogated using a range of sensitivity analyses. All analyses were undertaken using the TwoSampleMR R package.

To better understand the complex role of neutrophils in health and disease, we applied phenome-wide MR analysis (MR-PheWAS) to evaluate the consequences of genetically proxied variation in neutrophil traits on 14,983 outcomes including diseases, anthropometric and behavioral measures, plus blood concentrations of biomarkers, proteins and metabolites (Figure 1B). We leveraged GWAS summary data for three different neutrophil characteristics: 1) circulating blood neutrophil count (NEUT), 2) neutrophil side scatter (SSC), a measure of intracellular granule content, and 3) serum concentration of MPO, the most abundant neutrophil granule protein released upon cell activation.(9)

## MATERIALS AND METHODS

### Data sources

Analyses were undertaken using data from the largest European-ancestry GWAS for NEUT (n=170,702) and SSC (n=39,636) measured as part of the complete blood count test, and MPO (n=21,758) measured using the Olink proteomics platform (Table 1B).(10–12)

### Heritability and genetic correlation

SNP-based heritability and genetic correlation were calculated using linkage disequilibrium (LD) score regression (LDSC) software v1.0.1 with European LD scores generated from the 1000 Genomes Project reference panel.(13)

### Genetic instruments for neutrophil traits

For NEUT and MPO, all GWAS-significant single nucleotide polymorphisms (SNPs; p-value <5 ×10^−8^) were selected as potential instrumental variables (IVs). SNPs were clumped using an r^2^ threshold of 0.001 (within a 10,000kB window), resulting in a set of independently associated SNPs for each trait. For SSC, independent SNPs were selected using a multiple stepwise conditional analysis algorithm, as described previously.(11)

### Outcome data

Outcome traits were identified using the Medical Research Council Integrative Epidemiology Unit (MRC-IEU) open GWAS database, accessed via the R package TwoSample MR.(14) There were 50,044 traits with accessible GWAS summary data on 03/11/23. Filtering of traits was applied to exclude studies from participants of non-European ancestry, studies of gene expression and administrative codes (such as operative procedures) and any studies missing essential fields required to run MR analysis. This yielded 20,701 outcomes for analysis (Figure 1C).

### MR analysis

Analyses were undertaken using the TwoSampleMR R package. For each neutrophil trait PheWAS, associations for SNPs used as instrumental variables were extracted from summary GWAS data for the filtered outcome phenotypes. If a SNP was not present in the outcome GWAS, data from a proxy SNP in high LD with the target SNP (r^2^ > 0.8) was used, or the SNP removed if no proxies were available. Data were harmonized to ensure the effect of the variant on the outcome and exposure was relative to the same allele. For palindromic SNPs, effect allele frequencies were used to resolve coding strand ambiguities. Steiger filtering was then undertaken to select only those instruments which explained more variance in the exposure than the outcome, thereby minimizing the influence of reverse causality on the analysis. Two sample MR was conducted for all outcomes with ≥3 SNPs proxying the trait, using multiplicative random effects inverse variance weighed (IVW) estimates for the primary analysis. Results are expressed as the standard deviation change (SD, for continuous outcomes) or odds ratio (OR, for categorical outcomes) per SD increase in genetically predicted neutrophil trait, with corresponding 95% confidence intervals.

### Sensitivity analyses

MR p-values were adjusted for multiple testing using the FDR correction. Effect estimates using MR methods robust to pleiotropy (MR Egger, weighted median and weighted mode) were examined and Cochran’s Q statistic was used to test for heterogeneity in genetic instruments. MR analysis was also run in the reverse direction to examine the direction of causality for key findings.

## RESULTS AND DISCUSSION

We selected NEUT, SSC and MPO as exposure traits because they are relevant to neutrophil biology and can be measured using high throughput laboratory analyses of large population collections, thereby enabling robust genetic proxies to be determined by GWAS. SNP-based heritability for all three exposures were comparable to values reported for other blood cell traits (Figure 2A).(10)

**Figure 2:**
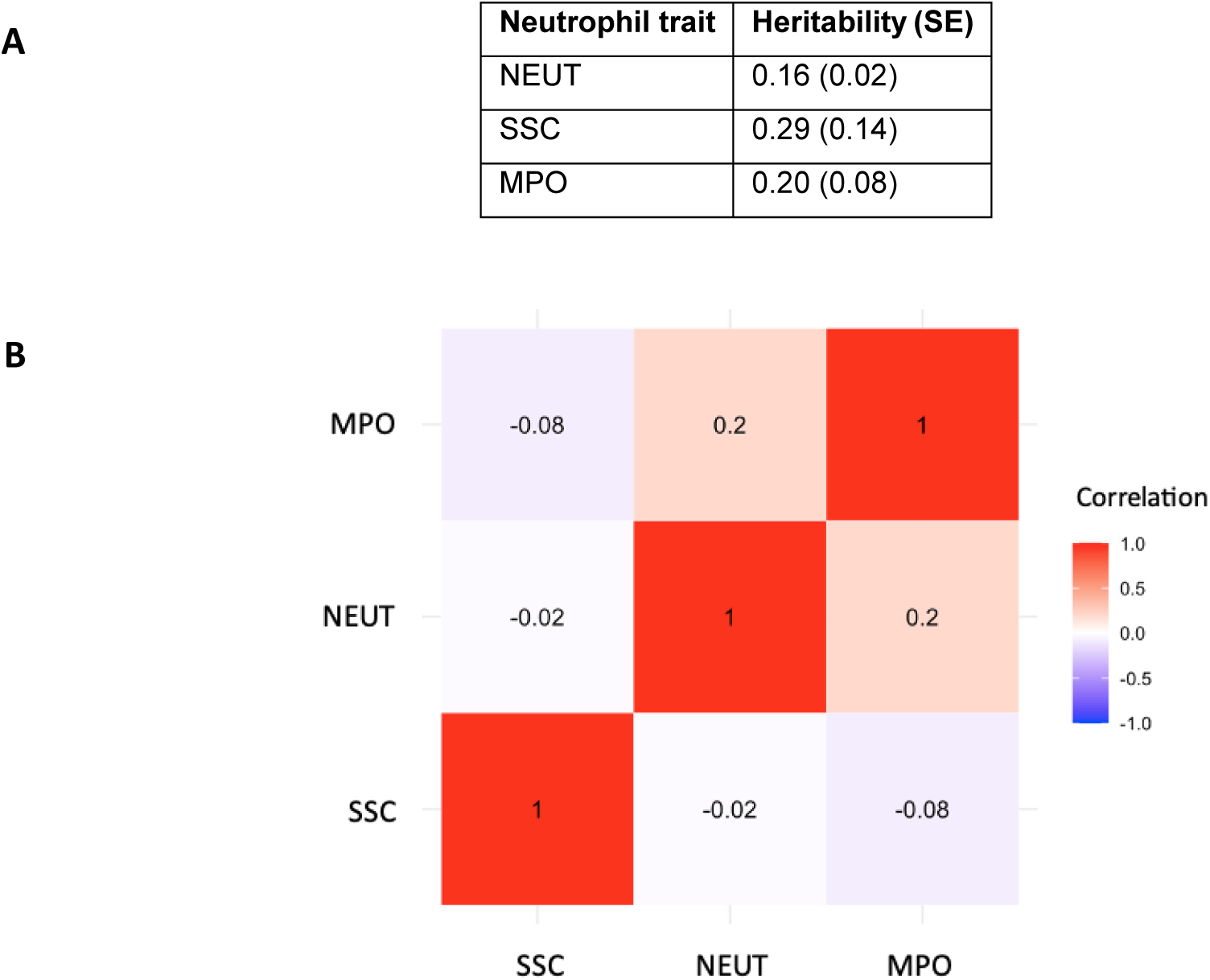
Heritability and genetic correlations for neutrophil traits. (A) Heritability for each neutrophil trait calculated using LD score regression. (B) Genetic correlation between neutrophil traits. Genetic correlations (rg) calculated using bivariate LD score regression.

Using independently associated SNPs from the largest published GWAS for each trait, NEUT, SSC and MPO were genetically proxied by 154, 72 and 10 variants, respectively. The mean F statistic, an estimate of the strength of each instrument-exposure association, was 74.7 for NEUT, 247.1 for SSC and 89.2 for MPO, thereby exceeding the widely reported threshold of >10 and indicating minimal risk of weak instrument bias for MR analyses.(15)

MR-PheWAS demonstrated that neutrophil traits had a genetically predicted effect on a total of 146 outcomes (FDR adjusted p-value <0.05; Table S1-3). Reflecting the number of SNPs used to instrument each trait, NEUT had the greatest number of associations (118), followed by SSC (26) and MPO (2; Figure 3).

**Figure 3.**
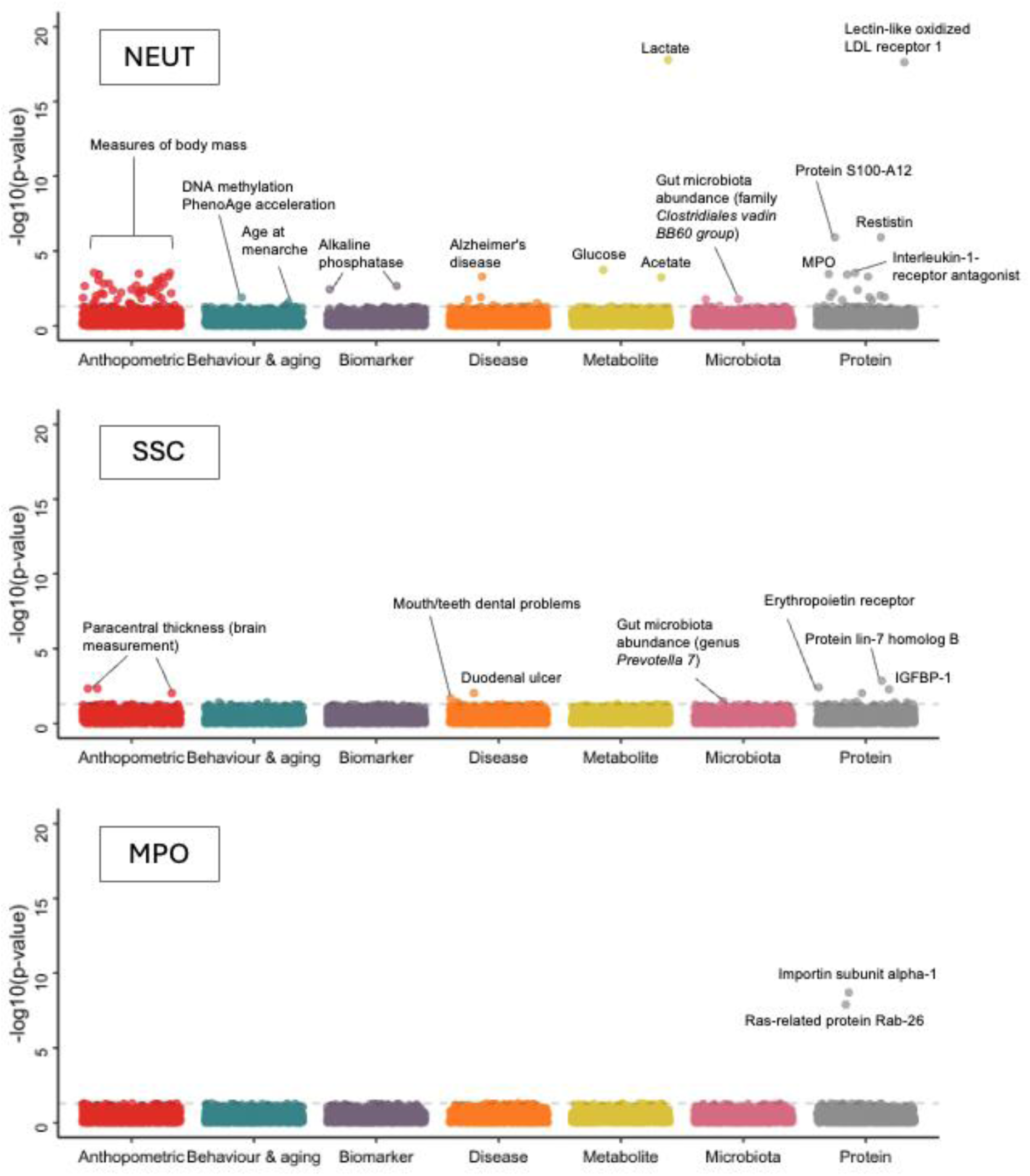
Phenome-wide associations with neutrophil characteristics. Manhattan plots for (A) NEUT, (B) SSC and (C) MPO. Hematological parameters excluded to aid visualization. Dashed grey line indicates FDR adjusted p-value of 0.05.

### Relationship between neutrophil count and blood cell phenotypes

Of the 118 associations with NEUT, 40 related to other blood cell parameters. These relationships were likely to be reflecting shared genetic architecture and/or polygenicity-induced horizontal pleiotropy, therefore were not explored in depth. However, we note a positive correlation between neutrophil count and quantification of all other blood cells, including RBC count. As neutrophilia is a widely reported marker of inflammation, this suggests that neutrophil count in population cohorts reflects increased haematopoiesis rather than chronic inflammatory states, typically characterised by anaemia. In keeping with this, genetically predicted increasing NEUT associated with higher serum alkaline phosphatase (ALP; SD 2.14 [1.16, 3.11], adjusted p-value 3.58 ×10^−3^), a measure of bone turnover, and there was little causal evidence for any of the neutrophil traits in this study on biomarkers of inflammation such as CRP or ESR (adjusted p-values >0.05).

### Neutrophil traits associate with plasma protein levels

The traits NEUT and MPO were noted to have a weak positive genetic correlation (rg 0.2), in keeping with previous functional studies (Figure 2B).(16, 17) Consistent with this, MR analysis identified an association between NEUT and MPO as well as seven other constituents of the neutrophil releasate including bactericidal permeability-increasing protein (Uniprot P17213; SD 0.30 [0.14, 0.44], adjusted p-value 1.92 ×10^−2^) and lactotransferrin (Uniprot P02788; SD 0.34 [0.18, 0.50], adjusted p-value 6.11 ×10^−3^). This supports the hypothesis that neutrophils have a basal release of neutrophil granules, therefore subjects with increased neutrophil counts have higher serum concentrations of granule markers.

SSC was not genetically correlated with either NEUT or MPO (rg −0.02 and −0.08 respectively), indicating that these are discrete cellular phenotypes. Both SSC and MPO associated with the serum concentrations of proteins implicated in intracellular membrane trafficking, including glycolipid transfer protein (Uniprot Q9NZD2; SD - 0.12 [−0.18, −0.07], adjusted p-value 9.54 ×10^−3^), polyphosphoinositide phosphatase (Uniprot Q92562; SD −0.11 [−0.17, −0.06], adjusted p-value 3.64 ×10^−3^), and Ras-related protein Rab-26 (Uniprot Q9ULW5; SD 0.89 [0.64, 1.15], adjusted p-value 1.25 ×10^−8^).

### Neutrophil count associates with lower body mass

Higher NEUT was causally associated with reduced body weight (SD −0.10 [−0.14, - 0.05], adjusted p-value 1.54 ×10^−2^), reduced body mass index (SD −0.07 [−0.10, - 0.03], adjusted p-value 4.52 ×10^−2^) and lower risk of obesity (OR 0.74 [0.65, 0.86], adjusted p-value 9.63 ×10^−3^; Figure 4). Pleiotropy robust MR methods showed concordant effect sizes and no reverse associations between body weight, BMI and obesity with NEUT as the outcome (Tables S1-3, 5). NEUT also showed associations with impedance derived-anthropometric traits such as reduced trunk, arm and leg predicted and fat-free mass, although there was evidence of confounding by reverse causality for these traits. Cochran’s Q statistics suggested heterogeneity between the SNPs used to instrument all measures of body mass, indicating potential pleiotropic pathways (Table S4). These findings may be indicative of a more complex relationship between neutrophils and body mass as prior observations have indicated that circulating neutrophil counts are elevated in obesity and that this corrects in response to weight loss after bariatric surgery.(18, 19)

**Figure 4.**
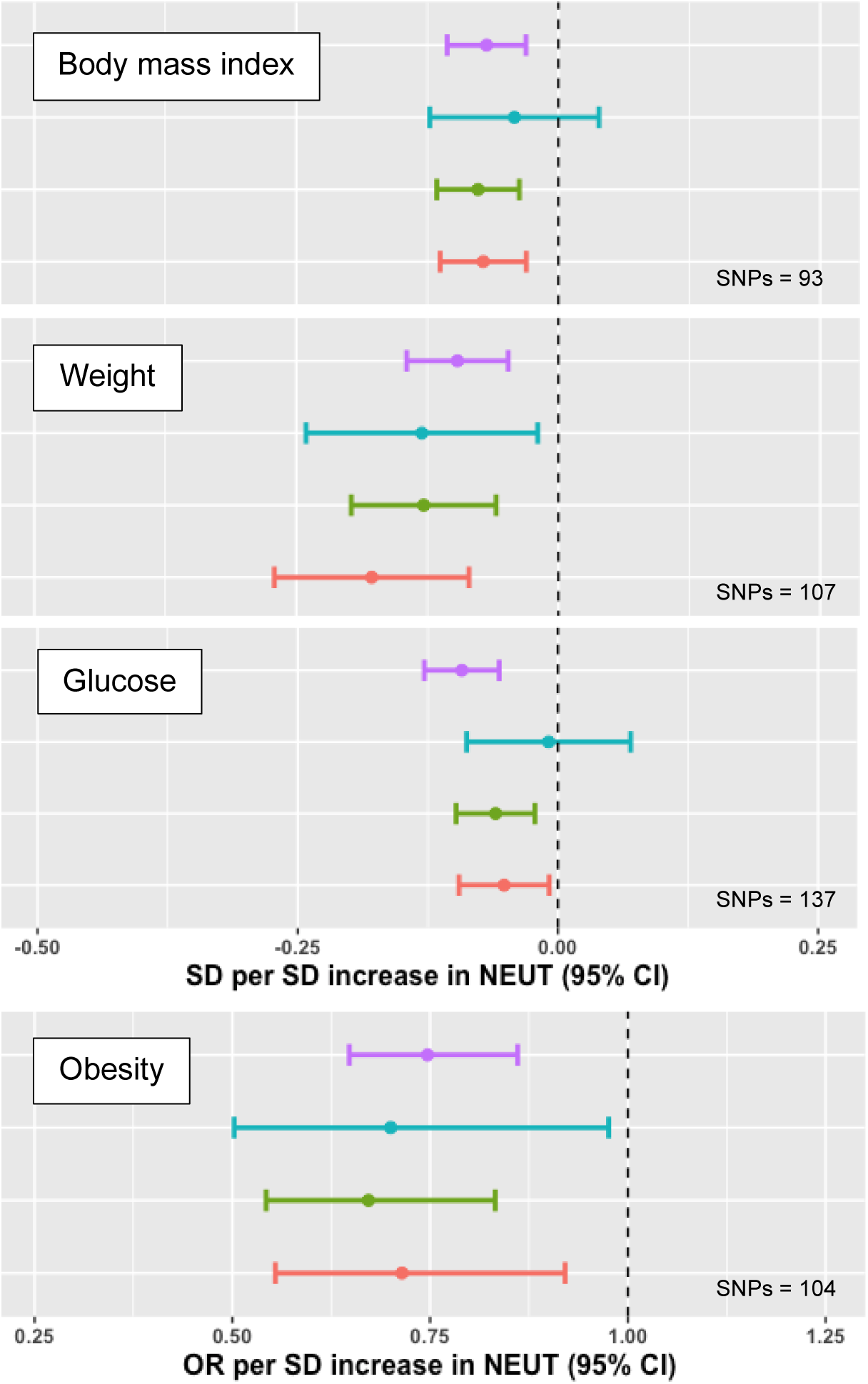
MR of neutrophil count associations with body mass. Forest plots with MR effect estimates for body mass index, weight, glucose and obesity. IVW (primary analysis) shown in in purple, MR Egger shown in blue, weighted median in green and weighted mode in red. Error bars signify 95% confidence intervals.

We identified several potential mechanisms by which neutrophils may reduce body mass. For example, increased NEUT was associated with lower serum glucose (SD −0.09 [−0.13, −0.06], adjusted p-value 1.75 ×10^−4^). Increased SSC associated with increased serum concentrations of both insulin-like growth factor-binding protein 1 (Uniprot P08833; SD 0.38 [0.22, 0.53], adjusted p-value 5.28 ×10^−3^), and angiopoietin-related protein 3 (Uniprot Q9Y5C1; SD 0.12 [0.06, 0.18], adjusted p-value 3.90 ×10^−2^) which are both implicated in regulating insulin sensitivity and have previously been inversely associated with obesity and metabolic syndrome, although no associations between SCC and measures of body mass were identified in our study.(20–23) Together, these findings suggest that neutrophils may influence body mass by altering glucose metabolism.

### Neutrophil count associates with endothelial activation but not with vascular disease

Increased NEUT associated with increased serum E-selectin (Uniprot P16581; SD 0.13 [0.08, 0.19], adjusted p-vale 5.01 ×10^−4^) and oxidized low-density lipoprotein receptor 1 (Uniprot P78380; SD 0.32 [0.16, 0.48], adjusted p-value 6.97 ×10^−5^), both of which are secreted by the vascular endothelium in response to inflammatory stimuli. This is in keeping with experimental findings that neutrophils promote endothelial activation.(24)

However, despite this association, we found little evidence for a causal relationship between any of the neutrophil traits and vascular pathologies such as hypertension, atherosclerosis, myocardial infarction, peripheral vascular disease or stroke (adjusted p-values >0.05). Multiple previous observational studies have shown that higher neutrophil counts are associated with increased risk of coronary heart disease.(25, 26) However, this finding was not replicated in our study, which accords with previous MR analyses (10, 27). This indicates potential confounding in the prior observational studies and highlights that genetic effects on intermediate phenotypes such as molecular pathways can be diluted or opposed downstream before exerting an influence on complex pathologies such as vascular disease.

### Neutrophil traits associate with alterations in the gut microbiome

MR analyses found that neutrophil traits were causally associated with alterations in the gut microbiome. Increased NEUT associated with the abundance of *Clostridiales vadin BB60 group* bacteria (SD 0.17 [0.08, 0.25], adjusted p-value 1.7 ×10^−2^) and increased neutrophil SSC associated with abundance of genus *Prevotella* 7 (SD 0.16 [0.08, 0.23], adjusted P = 3.25 ×10^−2^), both short chain fatty acid (SCFA) producing bacteria (Figure 5). In addition, NEUT was associated with reduced serum levels of acetate (SD −0.08 [−0.11, −0.04], adjusted p-value = 5.58 ×10^−4^), the most abundant gut-derived SFCA in the systemic circulation.(28)

**Figure 5.**
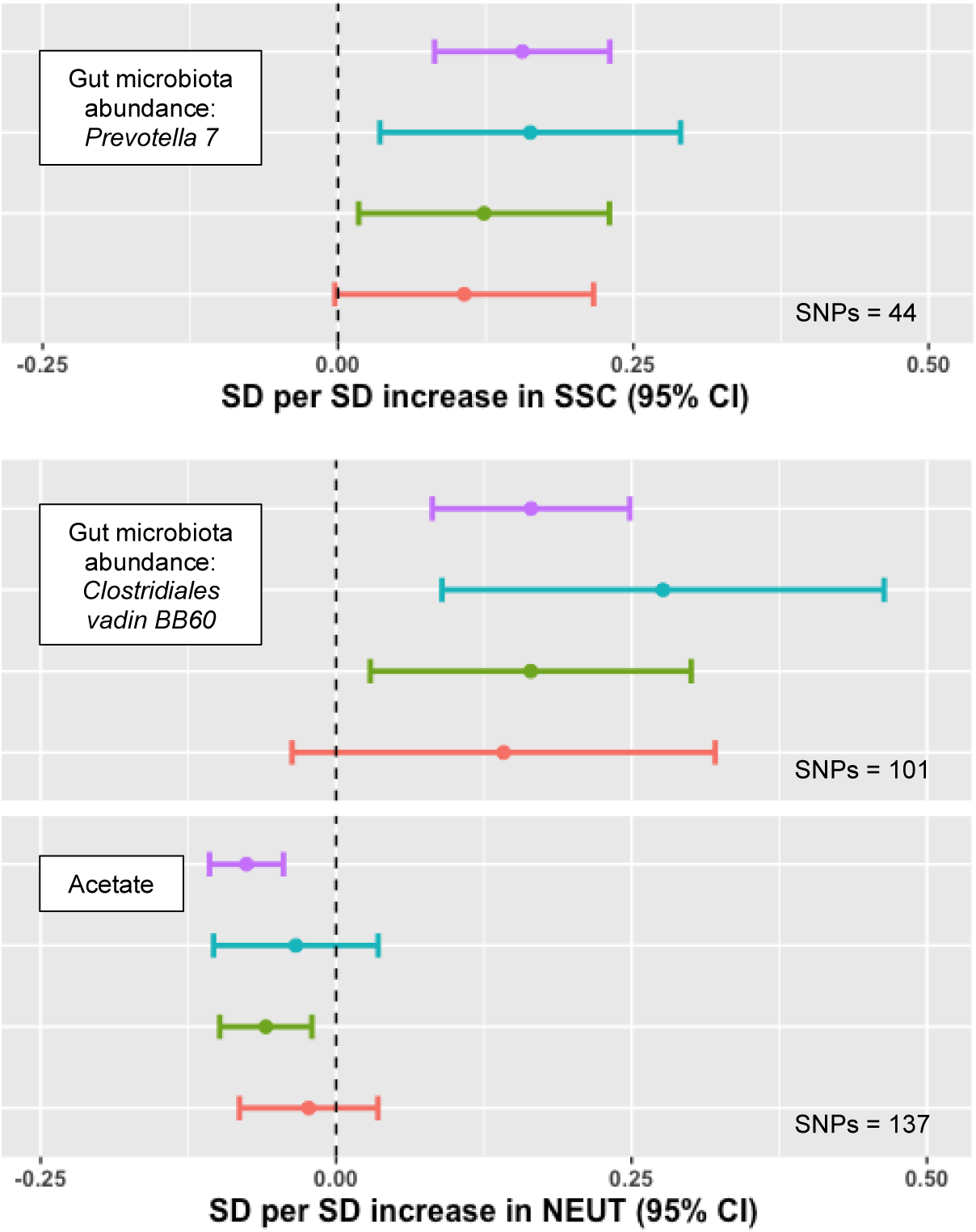
MR of neutrophil trait associations with the gut microbiome. Forest plots with MR effect estimates for abundance of gut microbiota and acetate weight. IVW (primary analysis) shown in in purple, MR Egger shown in blue, weighted median in green and weighted mode in red. Error bars signify 95% confidence intervals.

These findings are in keeping with the established role of neutrophils in regulating the composition of gut commensal species, with neutrophil dysfunction implicated in dysbiosis, an imbalance in microbial populations, which can promote disease states.(29, 30) Abundance of *Clostridiales vadin BB60* and *Prevotella* have been inversely associated with insulin resistance and obesity in previous observational studies, suggesting a further mechanism through which neutrophils may influence body mass.(31, 32) SSC was also associated with dental problems (OR 0.99 [0.99, 0.99], adjusted p-value 3.95 ×10^−2^) and duodenal ulcer (OR 0.99 [0.99, 0.99], adjusted p-value 9.54 ×10^−3^), both of which have been linked to alterations in gut microbiota, including *Prevotella,* in other studies.(33, 34) Interestingly, these phenotypes accord with the observation that patients with disorders of neutrophil number and/or function are prone to developing periodontitis and mucosal ulceration.(35, 36)

### Neutrophil count associated with risk of Alzheimer’s disease

We found that increased NEUT associated with an increased risk of Alzheimer’s disease (AD; OR 5.12 [2.62, 10.01], adjusted p-value 5.01 ×10^−4^; Figure 6). Pleiotropy robust MR estimates were concordant, Cochran’s Q statistic indicated homogeneity of IVs and MR in the reverse direction showed no significant associations, although we note that this association was proxied by only 3 SNPs. Increased SSC was associated with reduced measurements cortical thickness (SD - 0.09 [−1.10, −0.05], adjusted p-value 4.63 ×10^−3^), which is a proposed biomarker of neurodegeneration, correlating with cognitive impairment and development of AD.(37, 38)

**Figure 6.**
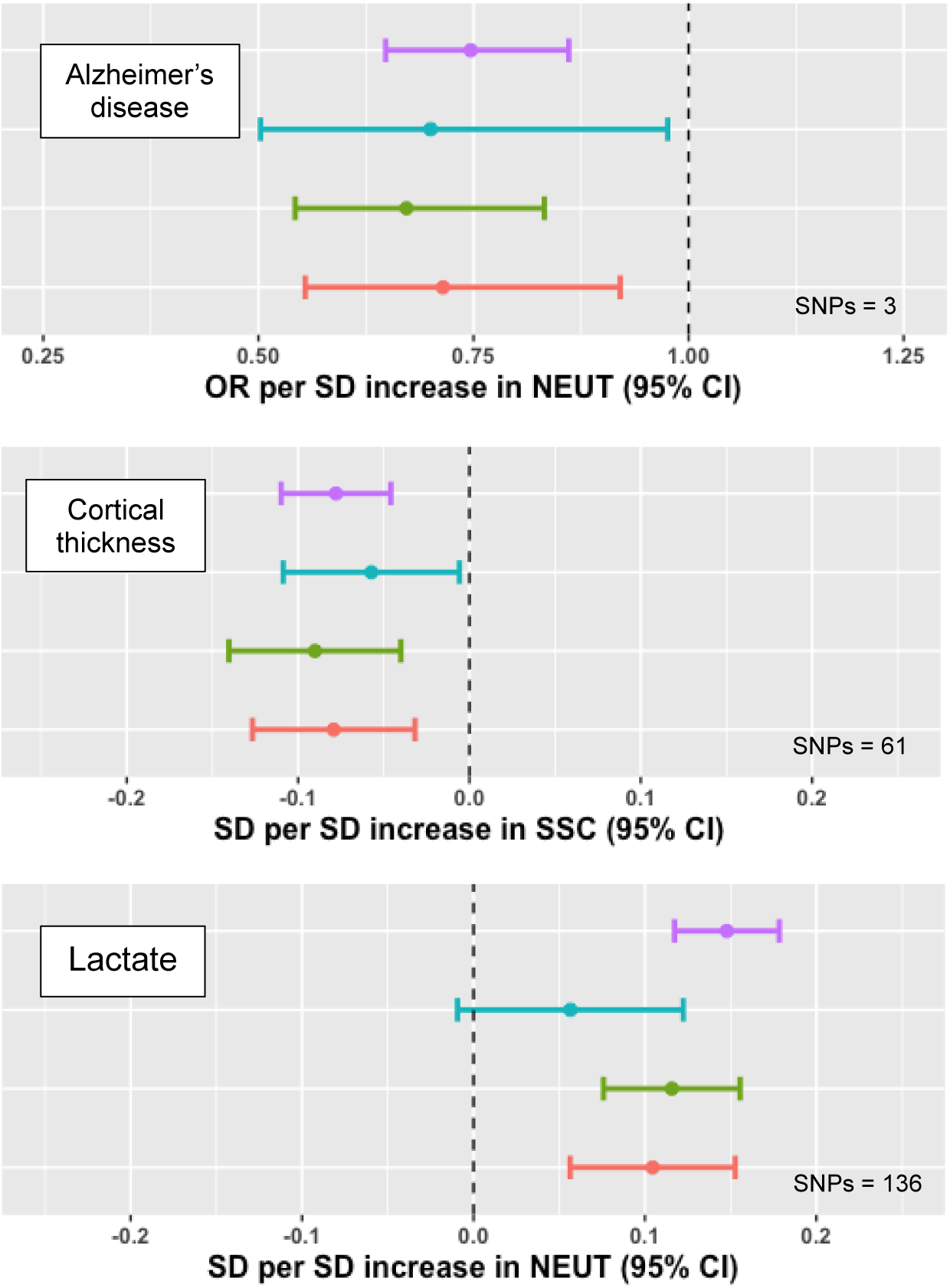
MR of neutrophil trait associations with Alzheimer’s disease. Forest plots with MR effect estimates for Alzheimer’s disease, cortical thickness and lactate. IVW (primary analysis) shown in in purple, MR Egger shown in blue, weighted median in green and weighted mode in red. Error bars signify 95% confidence intervals.

A possible role for neutrophils in the pathogenesis of AD has been suggested as neutrophil infiltration exacerbates neuroinflammation in animal AD models and neutrophil depletion or inhibition of recruitment reduces the formation of AD-associated amyloid-beta plaques.(39) Consistent with our findings, observational studies have linked neutrophil count to AD risk and reduced cognitive function.(40, 41) It is noteworthy that we also found that neutrophil count associated with blood lactate concentration (SD 0.15, [0.12, 0.18], adjusted p-value 1.64 ×10^−18^). Lactate, a metabolite produced by anaerobic metabolism in neutrophils, is a modulator of neuronal function and long-term memory and is increasingly implicated in AD pathogenesis.(42, 43)

### Study limitations

Although exposure IVs were constructed from the largest GWAS datasets available for the neutrophil traits, the power of our MR-PheWAS is constrained by the sample size of these studies, particularly for serum MPO concentration (n=21,758). Similarly, some disorders are either absent or only represented by small GWAS in MR-Base which was used to ascertain outcomes. There is likely to be some overlap between the exposure and outcome GWAS samples for some of the phenotypes, many of which included UK Biobank participants. This may potentially introduce bias leading to inflated type 1 error rates.(44) Finally, we acknowledge that our analysis was conducted using data from individuals of European ancestry and thus the results may not be generalizable to other populations.

### Conclusions

Here we present the first comprehensive MR-PheWAS for neutrophil traits and provide evidence for causal associations between neutrophil traits and outcomes relating to body mass, endothelial activation, the gut microbiome and Alzheimer’s disease. Hypotheses generated in this study now require orthogonal confirmation from observational population studies and focussed functional studies.

## Supporting information

STROBE-MR checklist

## Authorship Contributions

KF and KB designed the study and undertook data analysis. KF, KB and NC wrote the manuscript. AM, EV and BA supervised the study. All authors read, provided input and approved the final manuscript.

## Funding statement

This work was supported by the Wellcome Trust GW4 Clinical Academic Training Programme for Health Professionals to KF [227485/Z/23/Z] and NC [225541/Z/22/Z]. BA is funded by MRC grant MR/R02149X/1. KB is funded by an NIHR Academic Clinical Lectureship. For Open Access, the author has applied a CC BY public copyright license to any Author Accepted Manuscript arising from this submission.

## Acknowledgements

We acknowledge technical support from Ryan Langdon, Ruth Mitchell and Eleanor Sanderson (all University of Bristol).

## Data availability

R scripts used for the analysis are available via GitHub [https://github.com/kateburley/Neutrophil_PheWAS]. Harmonized summary data for all SNPs included in this analysis and full MR results are available through the University of Bristol Research Data Repository.

## Ethical approval

This analysis used GWAS summary data only, and therefore no additional ethical approval was required.

## Notes

**Conflicts of interest:** The authors declare no competing financial interests.

### Competing Interest Statement

The authors have declared no competing interest.

### Author Declarations

This analysis used GWAS summary data only, therefore no additional ethical approval was required. Approval was obtained by the original GWAS studies by their appropriate ethics review boards: see details in the source GWAS publications.

